# Anti-hypertensive Angiotensin II receptor blockers associated to mitigation of disease severity in elderly COVID-19 patients

**DOI:** 10.1101/2020.03.20.20039586

**Authors:** Yingxia Liu, Fengming Huang, Jun Xu, Penghui Yang, Yuhao Qin, Mengli Cao, Zhaoqin Wang, Xiaohe Li, Shaogeng Zhang, Lu Ye, Jingjun Lv, Jie Wei, Tuxiu Xie, Hong Gao, Kai-Feng Xu, Fusheng Wang, Lei Liu, Chengyu Jiang

## Abstract

**Background:** The novel coronavirus (CoV) severe acute respiratory syndrome (SARS)-CoV-2 outbreak started at the end of 2019 in Wuhan, China, and spread over 100 countries. SARS-CoV-2 uses the membrane protein Angiotensin I converting enzyme 2(ACE2) as a cell entry receptor. Indeed, it was reported that the balance of Renin-Angiotensin System (RAS), regulated by both ACE and ACE2, was altered in COVID-19 patients. It is controversial, however, whether commonly used anti-hypertensive drugs Angiotensin I converting enzyme inhibitor (ACEI) and Angiotensin II receptor blocker (ARB) shall be continued in the confirmed COVID-19 patients. This study was designed to investigate any difference in disease severity between COVID-19 patients with hypertension comorbidity. The included COVID-19 patients used ACEI, ARB, calcium channel blockers (CCB), beta blockers (BB), or thiazide to treat preexisting hypertension prior to the hospital were compared to patients who did not take any of those drugs.

**Methods:** In this multicentre retrospective study, clinical data of 511 COVID-19 patients were analyzed. Patients were categorized into six sub-groups of hypertension comorbidity based on treatment using one of anti-hypertension drugs (ACEI, ARB, CCB, BB, thiazide), or none. A meta-analysis was performed to evaluate the use of ACEI and ARB associated with pneumonia using published studies.

**Findings:** Among the elderly (age>65) COVID-19 patients with hypertension comorbidity, the risk of COVID-19-S (severe disease) was significantly decreased in patients who took ARB drugs prior to hospitalization compared to patients who took no drugs (OR=0·343, 95% CI 0·128-0·916, p=0·025). The meta-analysis showed that ARB use has positive effects associated with morbidity and mortality of pneumonia.

**Interpretation:** Elderly (age>65) COVID-19 patients with hypertension comorbidity who are taking ARB anti-hypertension drugs may be less likely to develop severe lung disease compared to patients who take no anti-hypertension drugs.

**Funding:** National Natural Science Foundation of China, Chinese Academy of Medical Sciences

**Research in context:** *Evidence before this study:* We searched PubMed for articles published up to March 15, 2020 using keywords “2019-nCoV”, “SARS-CoV-2”, “novel coronavirus”, and COVID-19 AND “ARB”, and “angiotensin II receptor blocker” for papers published in both English and Chinese. We found three papers: one from our group, published in Science China Life Science that demonstrated an elevated Angiotensin II level in blood samples from COVID-19 patients; another a perspective article in Chinese recommending ACEI and ARBs as potential remedies for SARS-CoV-2 infections; the third a retrospective study in Chinese identifying no significant difference between ACEI/ARB associated with outcomes in 112 COVID-19 patients with CVD comorbidity. The International society of Hypertension stated on March 16^th^, 2020: “there are no clinical data in human to show that ACE-inhibitors or ARBs either improve or worsen susceptibility to COVID-19 infection nor do they affect the outcomes of those infected”.

*Added value of this study:* We retrospectively reviewed different types of anti-hypertensive drugs taken by COVID-19 patients with hypertension comorbidity prior to entering the hospital. We discovered that ARB hypertensive drugs were associated with a decreased risk of severe disease in elderly (age>65) COVID-19 patients (OR=0·343, 95% CI 0·128-0·916, p=0·025), the first evidence of ARBs association to COVID-19 infections in human. We conducted a meta-analysis in the literature and found that ARB has positive effects associated with morbidity and mortality of pneumonia.

*Implications of all the available evidence:* ARB drugs are widely used in the population with hypertension. Treatments with ACEI and ARBs should be continuous according to medical guidelines. RCT trials of ARB associated with morbidity and mortality of SARS-CoV-2 infection are recommended in the future.

## Introduction

At the end of 2019, a cluster of lethal pneumonia cases was first reported in Wuhan, China. A SARS-CoV-like coronavirus in the respiratory tracts of patients was soon isolated and the viral genome sequenced; it was later named SARS-CoV-2^1-4^. In severe cases of COVID-19 patients developed acute respiratory distress syndrome (ARDS) and often died with multiorgan dysfunction syndrome (MODS). By March 11^th^ of 2020, the virus infection had spread to over 100 countries and SARS-CoV-2 infection was labeled a global pandemic by the World Health Organization^5^. Studies have shown that the SARS-CoV-2 shares the cell entry receptor as SARS-CoV: angiotensin I converting enzyme 2 (ACE2)^1,3,4,6-9^, a crucial negative regulator of renin-angiotensin system (RAS). Angiotensin I converting enzyme (ACE) and ACE2 are homologues with distinct important functions in RAS. ACE cleaves angiotensin I to generate angiotensin II, whereas ACE2 decreases the level of angiotensin II and negatively regulates RAS system^10-12^. RAS plays a crucial role in maintaining blood pressure homeostasis, fluid, and salt balance^13,14^. Anti-hypertensive drugs ACE inhibitor (ACEI) and ARB target RAS and have been used clinically for decades^15-18^ to treat millions of patients worldwide. Controversy about novel use of RAS blockers has been raised. By March 12^th^ 2020, the European Society of Hypertension (ESH) stated that “the currently available data on COVID 19 infections did not support a different use of RAS blockers”. The international society of Hypertension endorsed the ESH statement on March 16^th^ 2020 and stated “there are no clinical data in human to show that ACE-inhibitors or ARBs either improve or worsen susceptibility to COVID-19 infection nor do they affect the outcomes of those infected^24^”.

Our previous studies on SARS-CoV and acute lung failure elucidated the molecular pathogenesis signaling through RAS^19^. SARS-CoV infection and the spike protein of SARS-CoV reduce ACE2 expression, increase angiotensin II level signaling through the angiotensin II type1a receptor (AT1a), promote disease pathogenesis, induce lung edemas and impair respiratory function^8,11^. We demonstrated that losartan, a commonly used ARB drug, could attenuate acute lung failure in a mouse model whose condition had been worsened after injection of SARS-CoV spike protein^8^. We reported that RAS was imbalanced in the cases of many diverse predisposing factors for ARDS including sepsis, acid aspiration, bacteria, SARS-CoV, and avian influenza (H5N1 and H7N9) infections, as well as nanoparticle aspiration^8,10-12,20^. These studies indicated that blocking the RAS pathway and reducing angiotensin II level could ameliorate mice lung injury^8,10,11,15,20^.

Previous population-based studies of ACEI and ARB have shown use of the drugs to be largely associated with improved pneumonia outcomes^21^. Our previous study reported that plasma level of Angiotensin II in COVID-19 patients was significantly elevated^7^, indicating a altered RAS balance in COVID-19 patients. Here we conducted a retrospective study on the effects of RAS blockers on disease severity in COVID-19 patients. We also performed an updated meta-analysis of ACEI and ARB effects in reducing the risk of pneumonia and on pneumonia-related outcomes.

## Methods

### Study design and patients

Medical records of three cohorts (adult patients ≥18 years old) were included in this retrospective study: COVID-19 pneumonia admitted to the Shenzhen Third People’s hospital (Shenzhen, China) from Jan 11 to Feb 5, 2020; patients with COVID-19 admitted to the Renmin Hospital of Wuhan University (Wuhan, China) from Jan 12 to Feb 9, 2020; and patients with COVID-19 admitted to the Fifth Medical Center of People’s Liberation Army General Hospital (Beijing, China) from Dec 27, 2019 to Feb 29, 2020. All patients were diagnosed with COVID-19 pneumonia basing on the New Coronavirus Pneumonia Prevention and Control Program published by the National Health Commission of China^22^. Patients with hypertension comorbidity follow the criteria of National Guidelines for Hypertension Management in China (2019)^23^. This study was performed in accordance with guidelines approved by the Ethics Committees from the Institute of Basic Medical Sciences, Chinese Academy of Medical Sciences (002-2020). Verbal informed consent was obtained from patients or patients’ family members if available and the requirement for informed consent was waived by the Ethics Commission.

### Clinical data collection

Demographic, clinical, disease severity, and outcome data were extracted from medical records. Disease severity was classified as severe or mild according to the guidelines of 2019-nCoV infection from the National Health Commission of the People’s Republic of China. All data were independently checked by more than one physician.

### Meta-analysis

We searched the NCBI and Medline databases for potentially eligible records. Articles related to ACEI or ARB drug effects on pneumonia were considered in our meta-analysis. The search terms were as follows:

#1: pneumonia

#2: (angiotensin II Receptor Blocker) OR ARB

#3: irbesartan OR losartan OR valsartan OR telmisartan OR Candesartan OR Irbesartan

#4: (ace inhibitor) OR (angiotensin-converting enzyme inhibitor)

#5: captopril OR enalapril OR perindopril OR fosinopril OR lisinopril OR quinapril OR ramipril OR cilazapril OR benazepril

#6: #2 OR #3 OR #4 OR #5

#7: #1 AND #6

Non-human studies, non-English articles, articles not related to ARB or ACEI drugs effect on pneumonia, and case reports were excluded. The Newcastle-Ottawa Scale (NOS)^24^ was used for assessing the quality of studies included in meta-analysis by three researchers independently.

We collected the data from each article including journal name, publication year, first author’s name, population included in the article research, duration of follow-up, whether the study was prospective or retrospective, total sample size, treatment drug (ARB/ACEI and control), and risk results (OR) in the article.

The primary endpoint of articles included was defined as the incidence rate of pneumonia, and the secondary endpoint was the mortality of pneumonia patients. The diagnosis of pneumonia was based on clinical symptoms, chest radiograph results, and the criteria of International Classification of Diseases (ICD)^25^ provided in included articles.

### Statistical analysis

Mann Whitney U tests were used to compare differences in continuous variables between severe and mild COVID-19 pneumonia patients. Chi-squared tests were used to compare differences in categorical variables between severe and mild COVID-19 pneumonia patients. Odds ratio (OR) and 95% confidence interval (CI) were calculated for risk evaluation. Adjusted OR were calculated using “Enter” stepwise multivariable logistic regression^32^. Statistical analyses were performed with SPSS 16·0 for Windows (SPSS, Inc.). Chi-square tests were used to examine heterogeneity between trials; inverse variance (IV) method of fix or random effect model in meta-analysis were used based on the results of heterogeneity analysis. The software Review Manager 5 (RevMan 5) was used for meta-analysis. Two tailed P value<0·05 was considered to be statistically significant.

## Results

To evaluate the effects of RAS blockers on disease severity of COVID-19 patients, we conducted a retrospective study by obtaining hypertension comorbidity patient information from three cohorts and recording the types of anti-hypertensive drugs they took prior to SARS-CoV-2 infection. A total of 78 COVID-19 patients with hypertension comorbidity were qualified for the study (Figure 1). Among the 78 patients, 40 were classified as having mild disease (COVID-19-M), and 38 were classified as severe condition (COVID-19-S) according to New Coronavirus Pneumonia Prevention and Control Program published by the National Health Commission of China^22^. Age and sex were found to be statistically significant differences between COVID-19-M and COVID-19-S patients (Table S1)^7,26^. Our previous report showed that the AUC of ROC for age as a predictor of disease severity is 1^7^. There is no statistical difference in disease severity between any of the 5 different types of anti-hypertensive drugs (CCB, ARB, ACEI, Thiazide or BB) compared to no drugs taken by all COVID-19 patients with hypertension comorbidity in the study (Table S1). We further analyzed the data from 46 elderly COVID-19 patients over the age of 65 and found that patients who took ARB drugs prior to entering the hospital had a reduced risk of disease severity upon SARS-CoV-2 infection (OR=0·343, 95% CI 0·128-0·916, p=0·025) (Table 1). The effect of ARB remains positive in protecting from severe respiratory failure upon SARS-CoV-2 infection after the data are adjusted by multivariable logistic regression modeling with a sex variable (OR=0·25, 95% CI: 0·064-0·976, p=0·046) (Table 1).

**Table 1.** Association between antihypertensive use and disease severity of COVID-19 patients older than 65 years old with hypertension comorbidity.

| Characteristics |  | Total patients (n=46) | Severe patients (n=28) | Mild patients (n=18) | Unadjusted |  |  | Adjusted |  |  |
| --- | --- | --- | --- | --- | --- | --- | --- | --- | --- | --- |
|  |  |  |  |  | OR | 95% CI | p value | OR | 95% CI | p value |
| Antihypertensive use, n(%) |  |  |  |  |  |  |  |  |  |  |
|  | No use | 8 (17.4) | 7 (25) | 1 (5.6) | ref |  | ref | ref |  | ref |
|  | CCB | 26 (56.5) | 18 (64.3) | 8 (44.4) | 0.791 | 0.548-1.141 | 0.403 | 0.359 | 0.036-3.58 | 0.382 |
|  | ARB | 10 (21.7) | 3 (10.7) | 7 (38.9) | 0.343 | 0.128-0.916 | 0.025 | 0.250 | 0.064-0.976 | 0.046 |
|  | ACEI | 2 (4.3) | 1 (3.6) | 1 (5.6) | 0.571 | 0.139-2.342 | 0.378 | — | — | — |
|  | Thiazide | 3 (6.5) | 0 (0) | 3 (16.7) | — | — | — | — | — | — |
|  | BB | 7 (15.2) | 3 (10.7) | 4 (22.2) | 0.49 | 0.2-1.198 | 0.119 | — | — | — |

ARB, angiotensin receptor blocker; ACEI, angiotensin converting enzyme inhibitor; CCB, calcium channel blocker; BB, beta blocker.
OR, odds ratio; CI, confidence interval.
Adjustment was by multivariable logistic regression modeling with sex variable

**Figure 1.**
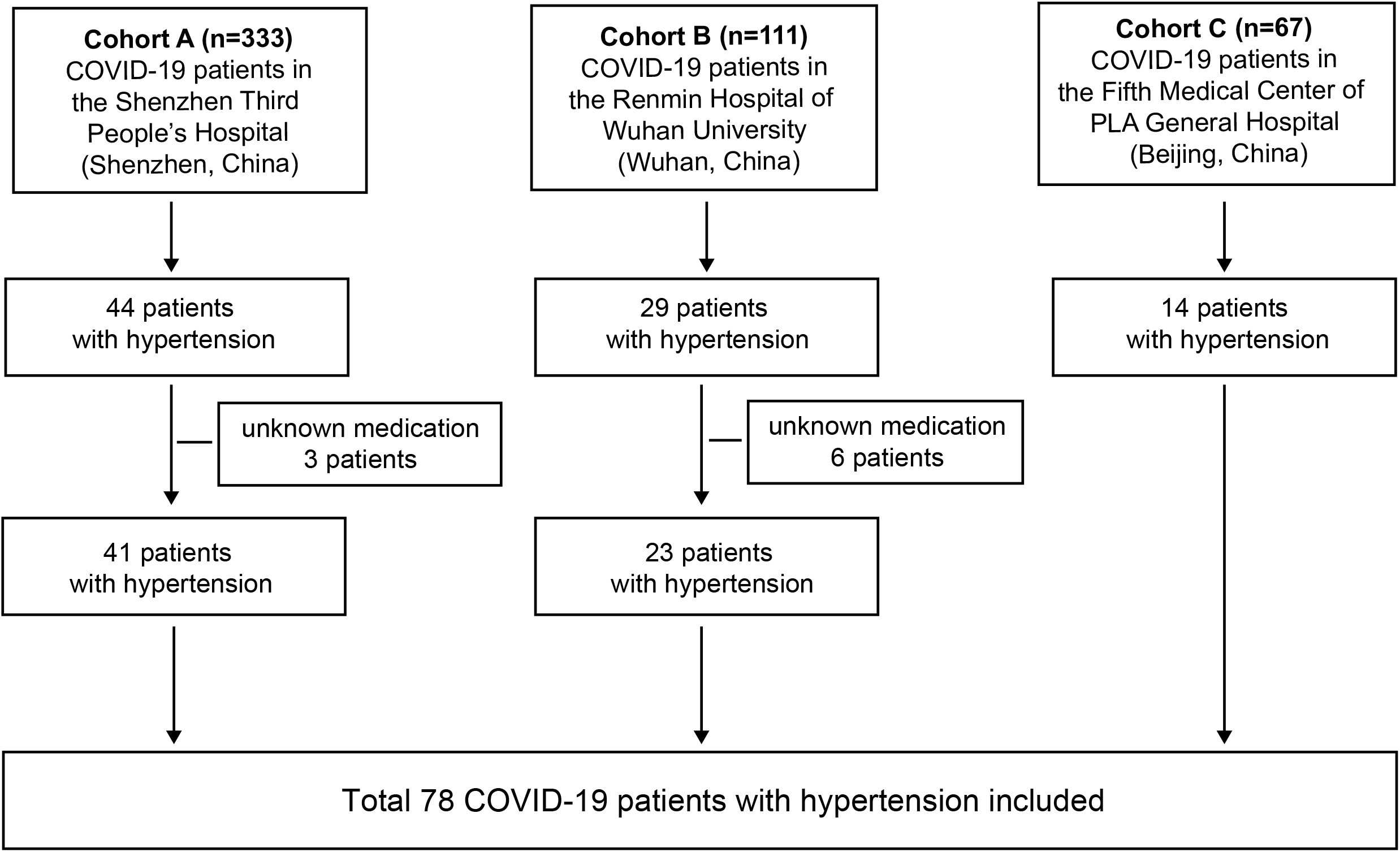
Study design and included patients. A total of three cohorts (adult patients ≥ 18 years old) were included in this retrospective study. Cohort A recorded 333 COVID-19 pneumonia patients admitted to the Shenzhen Third People’s hospital (Shenzhen, China) from Jan 11 to Feb 5, 2020; Cohort B recorded 111 COVID-19 pneumonia patients admitted to the Renmin Hospital of Wuhan University (Wuhan, China) from Jan 12 to Feb 9, 2020; Cohort C recorded 67 COVID-19 pneumonia patients admitted to the Fifth Medical Center of People’s Liberation Army (PLA) General Hospital (Beijing, China) from Dec 27, 2019 to Feb 29, 2020. In total, 78 COVID-19 patients woith hypertension comorbidity (having a with antihypertension medication record) were included in this study.

Furthermore, we performed meta-analysis of ACEI and ARB drugs associated with morbidity and mortality of pneumonia. By searching the Pub-Med database, we found 210 potentially eligible published papers and included 31 studies (Figure S1, Table S2). There are few detailed studies in the literature concerning ARB drugs associated with pneumonia studies, however ARBs were found to be associated with the reduced risk of pneumonia morbidity in a total of 70346 patients in three studies (OR=0·55, 95% CI; 0·43-0·70, p<0·01) (Figure 2A). By analysis of OR values from three studies available in the literature, ARBs were found to be associated with a decreased mortality rate for pneumonia (OR=0·55, 95% CI; 0·44-0·69, p<0·01), but not morbidity (Figure 2B-C). Previous meta-analysis studies have reported that ACEI drugs reduced morbidity and mortality of pneumonia patients^27^. Our updated meta-analysis of ACEI confirmed these results (Figure S2, S3). Taken together, our results indicate that ARB anti-hypertensive drugs associated with reduced disease severity of elderly COVID-19 patients and may have positive effects on morbidity and mortality of all-cause pneumonia.

**Figure 2.**
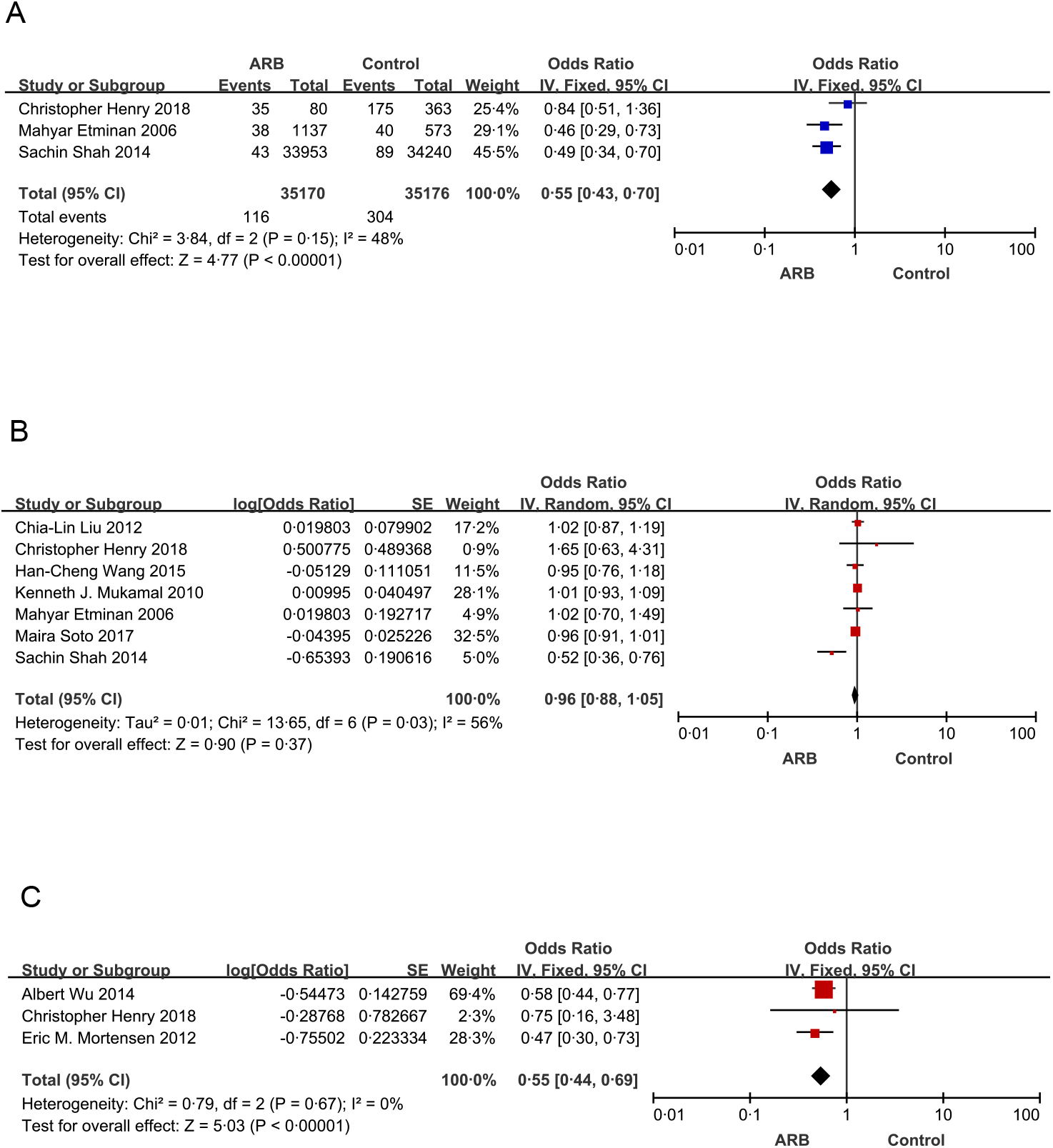
Meta-analyses of the pneumonia risk in patients taking ARBs. (A-B) Meta-analysis of the risk of morbidity of pneumonia in patients taking angiotensin II receptor blockers (ARBs) compared with no medication or other antihypertension treatment as control. (A) A total of 70346 patients in the three studies with detailed case/control numbers. (B) Seven studies with OR values. (C) Meta-analysis of the risk of mortality in pneumonia patients taking angiotensin II receptor blockers (ARBs) compared with no hypertension treatment as control. Three studies with OR values. CI confidence interval; IV, inverse variance; SE, standard error.

## Discussion

In this retrospective study, we report that ARB anti-hypertensive drugs significantly reduced the risk of severe pneumonia in elderly COVID-19 patients with hypertension comorbidity. There is no statistical difference in disease severity between patients who took CCB anti-hypertensive drugs compared to those who took no anti-hypertensive drugs (Table 1). Due to the limited number of COVID-19 patients who had taken ACEI, BB, and Thiazide prior to hospitalization, the association of these anti-hypertensive drugs with disease severity of COVID-19 patients with hypertension comorbidity had no statistical significance compared to patients who had taken no drugs. Further studies including more cohorts of COVID-19 patients are necessary in the future.

A previous retrospective study reported that ACEI and ARBs had no statistical significance in the outcomes of 112 COVID-19 patients with combined cardiovascular diseases (CVD) ^28^. The average age of the cohort in that study was relatively young at 62 years (range 55-67), and the disease severity of most cases was mild (96 of 112 cases)^28^.

Although the number of studies of ARB associated with pneumonia is limited in the literature, and none of the studies is a randomized controlled trial, our meta-analysis shows the positive effects of ARB associated with morbidity and mortality of pneumonia. Interestingly, none of the studies in the meta-analysis showed negative effects of ARB associated with morbidity and mortality of all-cause pneumonia.

The molecular mechanism of ARDS and severe pneumonia involved in RAS is complicated. Although the predisposing factors that induce ARDS are diverse (SARS-CoV, sepsis, acid aspiration, bacteria, and avian influenza (H5N1 and H7N9) infections, as well as nanoparticle aspiration), the imbalance of RAS seems to be a common mechanism^8,10-12,20^. Our previous studies have elucidated that ACE, angiotensin II, and the angiotensin II type 1a are problematic components in RAS that lead to disease pathogenesis, while ACE2 and the angiotensin II type 2a are beneficial components of RAS that protect mice from severe acute lung injury^8,11^. The level of angiotensin II in patient blood samples was usually elevated compared to healthy individuals^7,10,12^, and our previous study demonstrated that the plasma level of angiotensin II collected from COVID-19 patients was significantly increased^7^. Thus, the level of angiotensin II may indicate the RAS imbalance in patients. Regulators that balance RAS may be potential treatments of ARDS, and as such, ARB anti-hypertensive drugs might be an effective and economic remedy for COVID-19 patients already available in the market.

Moreover, it has not escaped our notice that ARBs may also have beneficial effects in the treatment of renal cancers, as well as heart failure ^29,30^. A study of the molecular mechanism underlying these treatment effects is necessary in the future. Taken together, our results not only support the continuous, wide use of ARBs in hypertension patients, but also suggest that an ARB trial for COVID-19-S patients in ICU with normal blood potassium level, normal renal function, normal blood pressure, or hypertension comorbidity is warranted.

## Data Availability

All the data in this study is available

## Contributors

CJ conceived the study concept and design. YL, MC, ZW, XL, and HG collected clinical data from medical records of 2019-nCoV infected patients admitted in Shenzhen Third People’s Hospital with LL’s supervision; JX, LY, JL, JW, and TX, collected clinical data from medical records of 2019-nCoV infected patients admitted in Renmin Hospital of Wuhan University; PY and SZ collected clinical data from medical records of 2019-nCoV infected patients admitted in The Fifth Medical Center of PLA General Hospital. FH led clinical data statistical analysis and meta-analysis with YQ assistants. KFX and FW provided critical help and discussion. CJ, FH, and YQ wrote the manuscript. The final version of manuscript was approved by all the authors.

## Acknowledgments

The authors would like to thank a scientist who insist to be anonymous. This work was supported by the National Natural Science Foundation of China (Grant 81788101), the Chinese Academy of Medical Sciences Innovation Fund for Medical Sciences (Grant 2017-12M-1-009), the Science and Technology Innovation Committee of Shenzhen Municipality (202002073000001), the National Key Research and Development Program (2020YFC0841700).

## Figure legends

**Figure S1. Literature search process for meta-analysis**.

NCBI and Medline databases were searched for potentially eligible records. Articles related to ACEI or ARB drugs effect on pneumonia were considered as potentially eligible studies. Non-human studies, non-English articles, articles not related to ARB or ACEI drugs effect on pneumonia, mixed data of ARB and ACEI drugs effect on pneumonia, and case reports were excluded. ACEI, angiotensin-converting enzyme inhibitor. ARB, angiotensin II receptor blocker.

**Figure S2. Meta-analyses of the pneumonia risk of patients taking ACEIs**.

(A) Meta-analysis of the risk of morbidity of pneumonia in patients taking angiotensin-converting enzyme inhibitors (ACEIs) compared with no medication or other antihypertension treatment as control. A total of 180493 patients in 16 studies with detailed case/control numbers were included. (B) Meta-analysis of the risk of mortality of pneumonia in patients taking angiotensin-converting enzyme inhibitors (ACEIs) compared with no medication or other antihypertension treatment as control. A total of 4084 patients in the three studies with detailed case/control numbers were included. CI confidence interval; IV, inverse variance; SE, standard error.

**Figure S3. Meta-analyses of the pneumonia risk of patients taking ACEIs (studies with OR values)**.

Meta-analysis of the risk of (A) morbidity (19 studies), and (B) mortality (8 studies) of pneumonia in patients taking angiotensin-converting enzyme inhibitors (ACEIs) compared with no medication or other antihypertension treatment as control. CI confidence interval; IV, inverse variance; SE, standard error.

